# Real-world datasets for the International Registry for Alzheimer’s Disease and Other Dementias (InRAD) and other registries: an international consensus

**DOI:** 10.1101/2025.01.12.25320418

**Authors:** Robert Perneczky, David Darby, Giovanni B. Frisoni, Robert Hyde, Takeshi Iwatsubo, Catherine J. Mummery, Kee Hyung Park, Johan van Beek, Wiesje M. van der Flier, Frank Jessen

## Abstract

**BACKGROUND:** Many dementia and Alzheimer’s disease (AD) registries operate at local or national levels without standardization or comprehensive real-world data (RWD) collection. This initiative sought to achieve consensus among experts on priority outcomes and measures for clinical practice in caring for patients with symptomatic AD, particularly in the mild cognitive impairment and mild to moderate dementia stages.

**OBJECTIVE:** The primary aim was to define a minimum dataset (MDS) and extended dataset (EDS) to collect RWD in the new International Registry for AD and Other Dementias (InRAD) and other AD registries. The MDS and EDS focus on informing routine clinical practice, covering relevant comorbidities and safety, and are designed to be easily integrated into existing data capture systems.

**METHODS AND RESULTS:** An international steering committee (ISC) of AD clinician experts lead the initiative. The first drafts of the MDS and EDS were developed based on a previous global inter-societal Delphi consensus on outcome measures for AD. Based on the ISC discussions, a survey was devised and sent to a wider stakeholder group. The ISC discussed the survey results, resulting in a consensus MDS and EDS covering: patient profile and demographics; lifestyle and anthropometrics; co-morbidities and diagnostics; imaging; treatment; clinical characterization; safety; discontinuation; laboratory tests; patient and care partner outcomes; and interface functionality.

**CONCLUSION:** By learning from successful examples in other clinical areas, addressing current limitations, and proactively enhancing data quality and analytical rigor, the InRAD registry will be a foundation to contribute to improving patient care and outcomes in neurodegenerative diseases.

## Introduction

The global landscape of dementia and Alzheimer’s disease (AD) registries is fragmented, with many operating at local or national levels without standardization or longitudinal comprehensive real-world data (RWD) collection [1]. This fragmentation limits advances in research and patient care. The Alzheimer Association’s ALZ-NET registry in the USA is a notable example of progress, collecting data on AD patients treated with novel FDA-approved therapies [2], highlighting the need for similar international efforts. By collecting comprehensive, harmonized data globally, an international registry would provide valuable insights into disease progression, treatment effectiveness and patient outcomes [3]. The registry would foster sustainable collaboration, promote knowledge sharing, facilitate patient recruitment for clinical trials, and help collect RWD for regulatory purposes [4]. Real-world evidence (RWE) complements randomized controlled trials by confirming their external validity in less-selected populations drawn from routine clinical practice [2-4]. Regulatory authorities increasingly mandate registry studies and RWD collection as part of risk management plans and other post-approval requirements [5-7] including, for example, biologics for chronic inflammatory diseases and monoclonal antibodies targeting amyloid-β [8, 9].

A RWD registry would provide evidence of treatment efficacy and safety outside of research settings, filling gaps left by controlled clinical trials [10]. An international registry would also help identify patients most likely to respond to specific treatments, including demographic measures, disease progression markers and genetic factors, enabling a more personalized approach to care [11]. Without a RWD registry, the necessary data are unlikely to be available from routine medical records as these also lack data collection standards and completeness. By tracking long-term patient outcomes in larger numbers and more diverse groups than is possible with clinical trials, registries can provide valuable insights into optimal care strategies, such as the best time to start or stop treatment, dosage adjustments, and managing side effects. Long-term tracking of patient outcomes will provide insights into optimal treatment strategies, and standardized biomarker assessments will facilitate early detection of AD, allowing for earlier intervention and more effective treatment [12].

Recent advances in interventions for early AD, particularly monoclonal antibodies targeting amyloid-β, have shown some efficacy and are being approved in different parts of the world [12]. These disease-modifying treatments (DMTs) slow disease progression and show positive results on different clinical and biomarker endpoints, including effective removal of amyloid-β from the brain as measured by positron-emission-tomography (PET) [13, 14]. As these drugs become more widely used, it will be crucial to explore how their efficacy in clinical trials translates to meaningful benefits for the patients and their safety profiles in real-world situations. Many questions on drug effectiveness and safety cannot be answered through controlled trials alone, and setting up registries for individual therapies is undesirable, underscoring the need for a global initiative to collect RWD from patients with and without different types and brands of treatment, ensuring that post-marketing monitoring of safety and effectiveness can effectively be performed in actual clinical care [15]. A global registry will help to set standards to improve the quality of care, facilitate the sharing of best practices, encourage the regular use of biomarkers in clinical practice, and help to avoid unnecessary treatments in patients unlikely to benefit [16].

The primary aim of this initiative was to define a minimum dataset (MDS) and extended dataset (EDS) to collect RWD in the new International Registry for AD and Other Dementias (InRAD) and across other AD registries globally. This effort sought to achieve consensus among experts on priority outcomes and measures for clinical practice in caring for patients with symptomatic AD, particularly in the mild cognitive impairment (MCI) and mild to moderate dementia stages. The MDS and EDS focus on informing routine clinical practice, covering relevant comorbidities and safety, and are designed to be easily integrated into existing data capture systems. The MDS and EDS document clinical characteristics and allow tracking of both existing and upcoming AD treatments, offering flexibility to include new outcome measures and novel therapies entering healthcare.

## Methods

### Definitions

In this paper, MDS refers to domains completed by every user in the context of diagnosis and management of AD, such as demographics, disease history, current conditions, and effectiveness and safety outcomes. EDS refers to data collected where practice permits, when additional instruments are used (eg those mandated by local authorities or that are part of local clinical practice).

### Organization of the consensus group

An international steering committee (ISC) of AD clinician experts was convened to lead the consensus initiative. Based on the expected approval of DMTs in 2025 in the EU, InRAD initially has a European focus, but the intention is to establish an RWD resource with global value. Hence, in addition to European experts, the ISC includes opinion leaders from countries with an approved DMT (Japan, UK) and non-EU countries with submitted DMT marketing applications (Australia, Switzerland). Countries with plans for adopting ALZ-NET were also considered (Republic of Korea). ISC members from other global regions, such as Africa and South America, will be added as the registry develops to ensure that countries are included that face unique healthcare challenges, demographic profiles, cultural factors, and resource availability. The overall aims and scope of the consensus were defined by this group and the process was managed by TW1 Healthcare Consulting Limited, London. The first drafts of the MDS and EDS were developed based on a previous global inter-societal Delphi consensus on outcome measures for AD [17] and additional data set established for the ALZ-NET project (www.alz-net.org/).

As a first step, a panel of participants was assembled (Table 1), representing key stakeholders who responded to a survey about the MDS and EDS. Stakeholders included clinical academics (N=72 out of 143 contacted individuals, response rate 50.4%), industry experts (N=4 companies) and patient representatives (N=4 organizations; one patient group provided two responses, and the weighting was adjusted to provide one response).

**Table 1.** Clinical, academic, patient representative and pharmaceutical company responses to the survey of the draft MDS and EDS.

| **Clinical and academic specialty** | **Number of responses (n=72)** |
| --- | --- |
| Neurologist | 51 |
| Geriatrician | 7 |
| Psychiatrist | 4 |
| Imaging specialist | 4 |
| Neuropsychologist | 4 |
| Genetics specialist | 1 |
| Remote and electronic data capture expert | 1 |
| **Patient representatives** | **Number of responses (n=5)** |
| Alzheimer's Association | 1 |
| Alzheimer's Disease International | 2* |
| Alzheimer Europe | 1 |
| Gates Ventures | 1 |
| **Pharmaceutical company representatives** | **Number of responses (n=4)** |
| Biogen | 1 |
| Eli Lilly & Company | 1 |
| Eisai | 1 |
| Roche Products | 1 |

\* Weighting adjusted to provide one response

Stakeholders from different geographical regions of the world were invited to respond to the survey to ensure that the consensus was relevant globally, including: 52 from Europe and Israel; 17 from Asia-Pacific Countries; and 3 from North America (Table 2). The academic group covered a range of diverse backgrounds, including: 51 neurologists; 7 geriatricians; 4 psychiatrists; 4 imaging specialists; 4 neuropsychologists; as well as a genetics specialist and an electronic data capture expert.

**Table 2.** Clinical and academic responses to the survey by country.

| **Country** | **Number of responses (n=72)** |
| --- | --- |
| Australia | 6 |
| Belgium | 1 |
| Canada | 1 |
| China | 1 |
| Denmark | 1 |
| Germany | 5 |
| Finland | 1 |
| France | 3 |
| Greece | 1 |
| Hong Kong | 1 |
| Iceland | 2 |
| Ireland | 2 |
| Israel | 2 |
| Italy | 1 |
| Netherlands | 4 |
| New Zealand | 1 |
| Norway | 3 |
| Portugal | 4 |
| Republic of Korea | 6 |
| Romania | 1 |
| Spain | 3 |
| Sweden | 2 |
| Switzerland | 4 |
| Taiwan | 2 |
| Turkey | 6 |
| United Kingdom | 6 |
| United States of America | 2 |

Members of the ISC and contributors to the stakeholder discussion and survey were not compensated for their work on the project. Eli Lilly and Company funded the involvement of TW1 Healthcare Consulting Limited, but did not have any influence over the consensus process outside of their inclusion in the stakeholder involvement.

### Consensus process

A meeting of the Steering Committee on 9 January 2024 discussed the first draft of the dataset. Based on these discussions, a survey was developed and sent to stakeholders between 9 and 26 February 2024. Survey respondents could designate each domain as MDS, EDS or ‘not applicable’ and comment on the frequency and/or measure used. Respondents could suggest additional domains and/ or outcomes for consideration by the ISC.

The survey results were discussed at ISC meetings held during the ADPD Conference in Lisbon on 5 March 2024 and on-line on 23 and 24 April 2024. Members who were unable to attend received copies of the minutes that they could comment upon. These comments were given the same weight as those from face-to-face and virtual attendees. The ISC discussed the safety and co-morbidity domains in detail during a series of virtual meetings between January and September 2024 and approved the final MDS and EDS in October 2024.

Consensus on each item was defined as at least 75% of participants agreeing that a domain should be in the MDS. Domains where 50% or fewer survey participants agreed were included in the EDS. The ISC discussed domains with 51% to 74% participants agreeing and decided whether the domain should be in the MDS or EDS. The survey included a ‘not relevant’ option, although this was not selected for any domain by any participant. The ISC also considered and discussed suggestions provided in open fields related to data elements, regional accessibility to scales, outcomes or markers, and considered the responses from participating pharmaceutical companies and other organizations.

Discordances that arose between pharmaceutical companies and patient representatives and the academic and clinician survey participants were used to direct the discussion where there was no concordance and to contextualize the discussion regarding the needs of the different stakeholder groups and how this data element should be included. This was especially relevant when the academic consensus was less than 75% but greater than 50%. In the context of this project the remit of the expert steering committee, who were selected due to their geographical representation and prominent track record in guiding and evolving the clinical practice in large dementia and memory clinics, was to evaluate the data elements that received a consensus score between 50 and 75% and to determine their validity as minimum data elements and adjudicated on the discrepancies in these grey areas. In addition, they also reviewed suggestions and recommendations from the survey and to consider their direct relevance to practice and whether the element should be included, adapted or adjusted for the data set. Of note, there are potentially multiple use cases for the registry data with the primary consideration for the minimum data elements being clinical care in the evolving setting of therapies impacting AD and considering the disparity and differences in current practice. If the disparity was for elements for specific research questions or elements challenging to collect in practice these data points were included in the EDS that would be potentially collected by interested centers or as part of registry studies or specific networks with common interest. Most importantly the possibility still exists for some minimum data elements to be recorded as not assessed for example where biomarkers or methods are not accessible or where ethical guidelines prohibit data collection in the case of diversity and sub-populations (e.g. ethnic background).

## Results

Informed consent will be mandatory and the availability of care partners and their willingness to be informants (assuming the patient agrees) will be ascertained. The ISC and consultation process resulted in a consensus MDS (Table 3) and EDS (Supplemental table 1). The domains are classified into several groups.

**Table 3.**
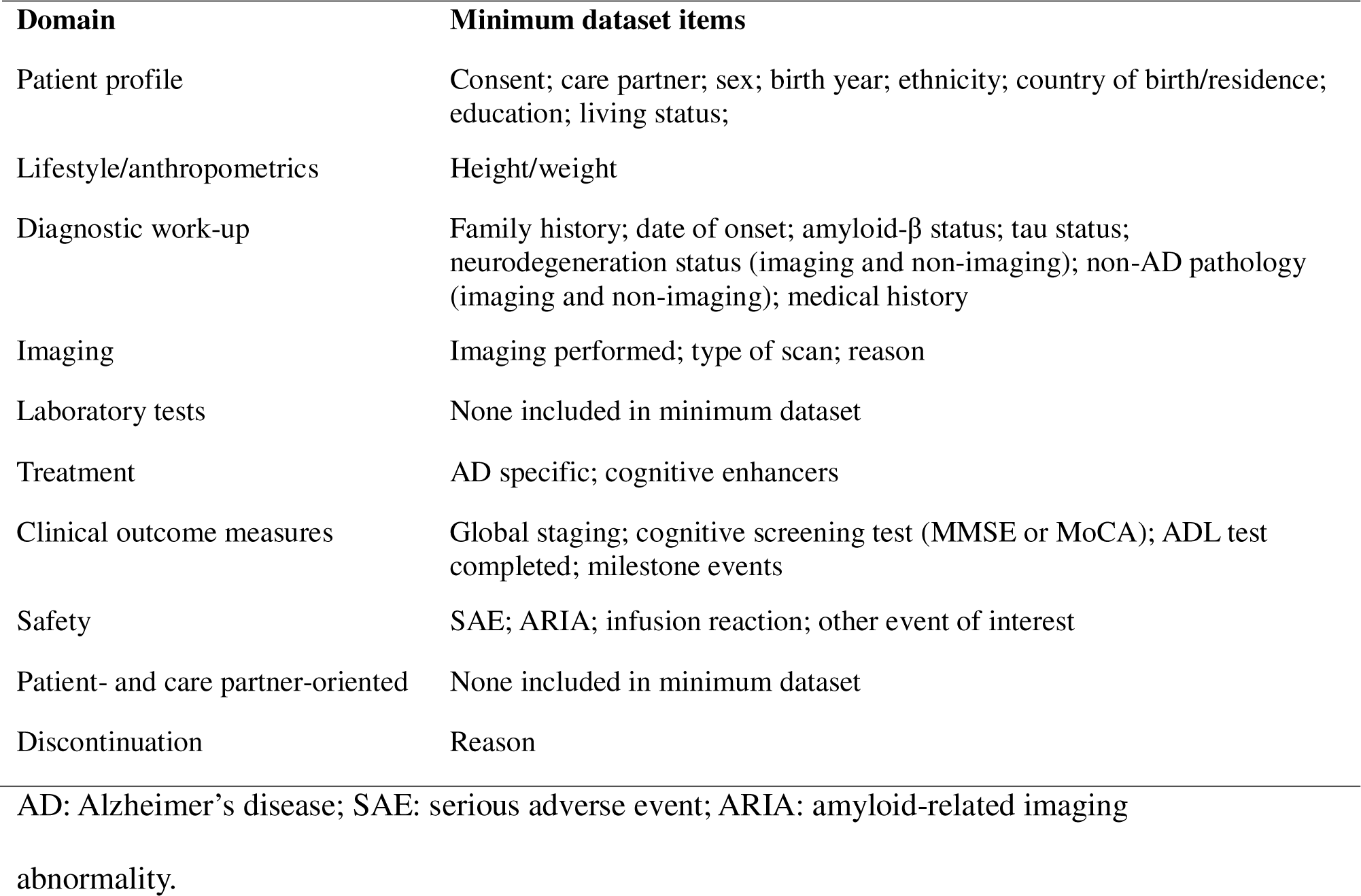
Domains identified by the International Steering Committee and items identified for the minimum dataset.

### Domain one: patient profile and demographics

The MDS includes eight subsections from this domain. Each patient will receive a unique ID. The EDS allows the user to include other patient identifiers most of which would be used in the local data set and defined as Personal Identifiable Information (PII). This data would be used at the local center and not uploaded to the registry database.

The MDS also includes basic demographics: sex assigned at birth; date/year of birth; race/ethnicity; country of residence; and education (assessed using the International Standard Classification of Education [18]). All eight sub-domains will be entered at entry (when permitted by local guidelines). The availability of care partners and their willingness to be informants may be updated at each visit however continuity is highly recommended. The EDS includes further domains that may be of practical value or facilitate deeper characterizations [19] including martial and work status, linguistic ability (eg bilingual) [20, 21] and dominant hand [22, 23].

### Domain two: lifestyle and anthropometrics

The MDS includes height (entry only) and weight (documented at entry and each clinic visit), which indicate body mass index [24]. The EDS includes further lifestyle domains: smoking, alcohol consumption, cannabis and recreational drug use, physical activity, active driving and sleep patterns [24-27].

### Domain three: Co-morbidities and diagnostics

The ISC acknowledged the impracticality of collecting every co-morbidity. Therefore, the ISC agreed on a list of relevant concomitant medical conditions to be recorded in the MDS that influence AD risk or modify the course of cognitive impairment or activities of daily living, such as cerebrovascular and other vascular or cardiac disorders, [28] psychiatric conditions [29], other neurological/neurodegenerative conditions (including traumatic brain injury) as well as other significant other historic or on-going conditions including metabolic conditions (eg diabetes, obesity), malignancies, blood or lymphatic diseases, immune or autoimmune conditions, or history of severe or recurrent infections [30]. The ISC agreed to capture medicines for concomitant diseases that potentially modify dementia outcomes (eg metformin) [31] as well as antithrombotic and antiplatelet medications, which have implications for current antibody therapies for AD [32] and which manage certain co-morbidities [33].

The patient’s history of these co-morbidities will be documented at entry to the registry. Current relevant co-morbidities will be updated at each clinic visit if ongoing. Whether there is a history of dementia in a first-degree relative, the date of the patient’s symptom onset, date of diagnosis, the syndromic presentation and etiological diagnosis (AD and co-morbidities), and the predominant clinical syndrome during the first two years will be recorded if known. In a case where the subject presents as asymptomatic or other syndromic presentation the changes in diagnosis can be added over time. In addition, the MDS will include as biomarker information the amyloid-β and tau-status, and whether there is evidence of neurodegeneration on imaging (see also domain group 4), with a ‘not performed’ option in the case where access to tests or imaging is limited.

The EDS includes further domains including details of the referral pathway, the specialty of the diagnosing physician and whether patients are or have been enrolled on a clinical trial. The EDS allows users to input the results of plasma neurofilament light chain measurements, and new biomarkers will be added as they become available.

### Domain four: Imaging

The MDS will include whether structural brain imaging was performed. These details will be input at entry and each clinic visit. The EDS contains detailed imaging-related domains including the scanner and magnet field strength, where relevant, information on atrophy, vascular lesion, and microbleeds, results of PET quantification and information on dopamine transporter imaging. Real-world brain scans and biomarkers present challenges for registries. Initially, a pragmatic project is planned to explore the feasibility of collecting the InRAD MDS and real-world imaging and fluid biomarker data across 12 memory clinic sites participating in the Dementias Platform UK (DPUK) Trials Delivery Framework (www.dementiasplatform.uk/trials-delivery/Trials-Delivery-Framework), which extends to 78 National Health Service (NHS) sites across the UK and includes sites with a high level of ethnic and socioeconomic diversity and deprivation. All 12 sites are organized in the Quantitative MRI in NHS Memory Clinics (QMIN-MC) network (https://www.rittman.uk/qminmc/; PI: T. Rittman) and use the NHS harmonized MRI protocol based on the UK Biobank MRI study [34]. A fully integrated, centralized data storage and analysis platform with a web-based portal for uploading and storing imaging data will be piloted in the feasibility study by a German technology partner (www.medotrax.com). An interface for standardized reporting will be implemented to support (neuro)radiologists in producing their reports. Automated quality assurance tools will empower sites to monitor and improve image acquisition and include analytics for image-based metrics. A research platform for sharing anonymized clinical and imaging data will be deployed, in collaboration with the DPUK imaging network, which has completed test-retest scans to support multi-site harmonization.

### Domain five: treatment

The MDS includes treatments prescribed for AD focusing on DMTs and symptomatic and other treatments of interest, including acetylcholinesterase inhibitors, memantine and nutritional supplements. The three MDS domains in this group will be collected at entry and updated at each visit. Moreover, the MDS includes the details (eg dose, route and frequency), and start, change and stop dates. Approved therapies will be presented as a drop-down list with international non-proprietary name (INN) and brand names. Other medicines will be entered as the INN rather than brand name. The list will be updated as new therapies become available. Non-pharmacological treatment options will be added later.

### Domain six: clinical characterization

The MDS includes four subsections from this domain - global, cognitive, functional, milestones - all of which will be collected at entry and updated at each visit (table 3). Global clinical staging will use the 2024 Alzheimer’s Association Workgroup revised criteria for diagnosis and staging of AD [35] based on clinical judgement to assign patients to one of six stages [35]:

- Stage 0: Asymptomatic, deterministic gene
- Stage 1: Asymptomatic, biomarker evidence only
- Stage 2: Transitional decline – mild detectable change, but minimal impact on daily function
- Stage 3: Cognitive impairment with early functional impact
- Stage 4: Dementia with mild functional impairment
- Stage 5: Dementia with moderate functional impairment
- Stage 6: Dementia with severe functional impairment

The ISC recommended that measures of cognitive function, such as the Mini-Mental Status Examination (MMSE) [17, 36, 37] and /or the Montreal Cognitive Assessment (MoCA) [17, 38, 39] should be part of the MDS. The clinician will determine whether the patient has attained a functional milestone. Given that activities of daily living scales are not part of the clinical routine assessment in most settings, the ISC does not recommend a specific instrument but rather agreed to monitor changes in three specific milestones (work status, driving status and living status) that impact on the life of the person with AD and their family.

Other assessments used in clinical practice can be entered into the EDS. In addition, as part of post-approval commitments, regulators may mandate completion of specific measures, such as the Clinical Dementia Rating Sum of Boxes (CDR-SB) [40, 41]. Therefore, CDR-SB is part of the EDS alongside other functional instruments such as the Neuropsychiatric Inventory Questionnaire (NPI*-*Q) [42] and the Utilization in Dementia*-* Lite (RUD-Lite) score [43, 44].

### Domain seven: safety

The MDS includes four subsections from this domain, all of which will be collected at each visit. The ISC acknowledged the impracticality of collecting every adverse event in clinical practice as the common events are known and documented in the label. The characterization of adverse events as related to a single therapeutic agent has also proved problematic in the real-world setting, and was raised in the survey, as subjects may be on or off therapy; hence the adoption of a universal term “Medical Events” has been recommended and is used in other disease registries. Medical events of special interest (MESI; table 3) are specific adverse events that are closely monitored due to their potential clinical significance or impact on patient safety. These events are identified based on their known or suspected association with a particular product, therapeutic class, or patient population. MESIs are predefined and rigorously tracked during clinical trials and post-marketing surveillance to detect any safety signal, and they segregate into two main groups: (1) MESI related to AD therapy (eg ARIA and infusion/injection reactions, including hypersensitivity) and (2) emergent MESIs that may have a direct or immediate impact on the patient’s functional status including serious malignancy, other neurological conditions, or fulfilling the criteria for a serious adverse event (SAE). A SAE results in death, is life-threatening, requires hospitalization or prolongation of existing hospitalization, results in persistent or significant disability or incapacity, or is a birth defect [45].

All ARIA [46] are included in the MDS, as required in registries on antibodies targeting amyloid-β [8] with management strategy (eg drug discontinuation, treatment resumption and outcome). In addition, the MDS includes infusion and injection reactions; and other adverse events that the ISC agreed were of interest. Additional events (eg hospitalization, out-patient, rehabilitation, treatment regimen modification) can be entered into the EDS.

### Domain eight: discontinuation

At the final visit at which the patient leaves the registry, the clinician will record the reason for discontinuation, such as death or withdrawal of consent [47]. It is important to note that patients who leave the study for reasons unrelated to death may do so because of worsening health or treatment-related issues. This could introduce a bias, as those who remain in the study may be systematically healthier. Without careful monitoring of those who discontinue for reasons other than death, there is a risk of overestimating the health of dementia patients, as the remaining participants may represent a subset of individuals who are in better condition, thereby skewing the overall findings. We will address this potential bias by including a plan for monitoring and analyzing the reasons for study discontinuation, ensuring that we account for the health status of those who leave the study for reasons other than death.

### Domain nine: laboratory tests

The laboratory test information for the EDS includes the biomarkers amyloid-β, tau, ptau and additional biomarkers as well as genetic information (*APOE* genotype) plus whether any material was biobanked and the assay used. In addition, standard blood chemistry and haematology fields will be available if needed.

### Domain ten: patient and care partner outcomes

The following measures of patient and care partner assessments are included in the EDS: the Quality of Life in AD scale (QoL*-*AD) [48, 49], Euro-QoL 5 dimensions 5 levels (EQ-5D-5L) [50, 51], the Dependence Scale [52], the Zarit Burden Interview [53, 54], the Amsterdam Instrumental Activities of Daily Living Questionnaire (A-IADL-Q) [55-57], and the AD Cooperative Study - Activities of Daily Living Scale for use in MCI (ADSC_ADL-MCI) [58].

### Interface functionality

The ISC agreed that the interface capturing the MDS and EDS will use a combination of drop-down menus, yes/no boxes and open-text fields. Figure 1 shows an example of the data-tree structure. As an example, living status (eg caregiver involvement) is entered at first visit. The healthcare professional checks whether there has been a change at each visit using yes/no boxes. If yes, the change is noted using a standard drop-down list and an open-text field that allows the inclusion of additional information. The data set includes a drop-down menu of ethnicity, with an open-text field for additional information.

**Figure 1:** Example of data tree structure. In this example, serious adverse events (SAE), amyloid-related imaging abnormality (ARIA) events and infusion/injection reactions since the last visit are checked by the healthcare professional using yes/no boxes. If yes is recorded for any of these events, the change is noted using a standard drop-down list and an open-text field that allows the inclusion of additional information. For the ARIA example this includes the start and stop date of the event, if symptoms were present or absent (if symptomatic the MedDRA code, severity and outcome are also recorded) and the relationship to the drug and action taken.

The ICS recommended that a prompt should remind users to complete specific items at each visit to ensure rapid and complete follow up with a time stamped denominator in all subjects over time. Each event occurrence should be described using standardized coding (eg Medical Dictionary for Regulatory Activities; MedDRA). The ISC also advocated prompts for treatment changes and co-morbidities, including documenting no change, which is considered as important as a change when using the registry to assess risk and disease progression.

## Discussion

RWD registries aim to bridge the gap between clinical trials and everyday clinical practice, providing comprehensive datasets reflecting the diverse patient populations and treatment scenarios encountered in real-world settings. We propose a registry-agnostic, internationally agreed MDS and EDS to collect practice-based RWD in AD of data arising in routine care, including patient history, clinical and biomarker outcomes, and safety profiles of drugs.

These datasets were composed by a consensus process including a diverse group of key stakeholders, aiming to create a common base while being sensitive to local, national and international variations in access to technology (eg neuroimaging [59, 60] and other biomarkers), measures in practice, such as the preferred cognitive assessment, clinician time constraints and existing or upcoming registries. This process resulted in a unified and internationally acceptable standard for RWD collection to support the evolution of AD care and to improve the study of the natural disease course and intervention effects.

The definition of the MDS and EDS was coordinated by InRAD (www.inradnetwork.org), a new interactive, dynamic, multinational effort to collect, organize, and provide feedback based on data collected in routine clinical practice. InRAD will use the datasets to capture and report longitudinal individual and aggregated data, with the launch in 2025 of a cloud-based, secure platform for data storage, sharing and analysis. The registry will be managed by a non-profit organization (InRAD Foundation) with an independent scientific leadership group, allowing partner sites to retain full ownership and control over their data. InRAD will be free to use and provide the participating clinicians with useful information about their patients at the point-of-care to facilitate patient counselling and education, and track and present changes in outcomes.

InRAD is distinguished from other large prospective AD data collections, such as ALZ-NET [61], and the National Alzheimer’s Coordinating Center (NACC) [62], through its unique focus on integrating RWD from diverse clinical settings. While ALZ-NET is specifically designed to collect RWE on the effectiveness and side effects of new FDA-approved AD treatments, and NACC provides a comprehensive dataset from multiple AD Research Centers across the US, InRAD emphasizes a broader, more inclusive approach, capturing data from a wide array of healthcare environments. This inclusivity allows InRAD to provide a representative picture of AD management in everyday clinical practice, leading to generalizable findings for application to improving patient care.

Integrating RWD from heterogeneous healthcare systems presents significant technological and logistical challenges, particularly in low-resource settings. One major challenge is ensuring data quality and reliability, as RWD comes from various sources with differing data collection standards and methods, leading to potential inconsistencies. Additionally, the standardization of data across diverse systems is complex, requiring robust protocols to harmonize data for meaningful analysis. Technological barriers, such as limited access to advanced data management tools and infrastructure, further complicate integration efforts in low-resource settings. To address these issues, we propose leveraging advancements in data analytics and artificial intelligence to enhance data standardization and integration. Furthermore, fostering collaborations with local healthcare providers and investing in capacity-building initiatives can help overcome logistical challenges and ensure the successful integration of RWD across diverse healthcare environments. By addressing these challenges, we aim to create a more comprehensive and reliable RWD repository that can inform and improve clinical practices globally.

The utility of RWD has been well-documented in other medical fields, particularly in oncology, cardiology, diabetes, and rheumatology. In oncology, RWD plays an important role in drug development, present across all development stages. For instance, a review of cancer drugs authorized by the European Medicines Agency (EMA) in 2018-2019 indicated that RWD was used 100% in discovery, 37.5% in early development, 58.3% in clinical development, 62.5% in registration decision and 100% in post-authorization lifecycle management [63]. RWD has also significantly impacted clinical practice, for example by helping to identify real-world endpoints and measure treatment outcomes in metastatic non-small cell lung cancer [64]. In cardiology, registries like the American College of Cardiology’s National Cardiovascular Data Registry (NCDR) [65] have significantly contributed to improving cardiovascular care. These registries collect data on various cardiovascular conditions and procedures, helping to identify best practices and improve patient outcomes [66]. RWD has been used to understand the effectiveness, safety, and costs associated with treatment options, complementing clinical trial findings and filling knowledge gaps.

In the field of diabetes, RWD have been pivotal in improving the prevention and management of diabetes-related outcomes. The PIONEER REAL Switzerland study [67], for example, provided valuable insights into the effectiveness of oral semaglutide in routine clinical practice, showing significant reductions in glycated hemoglobin (HbA1c) and body weight among participants, supporting the drug’s use in real-world settings. RWD is also the foundation of studying the effectiveness of semaglutide in AD [68, 69]. RWD are also widely collected in MS. InRAD is based on well-established secure, two-system platform used in MS (MSBase). Sub-studies using MSBase helped investigate diverse, but clinically important issues, such as: disease activity in pregnant and postpartum women receiving DMTs [70]; DMT prescribing patterns during the COVID-19 pandemic [71]; and routine CSF parameters as predictors of disease course [72]. Despite the promising potential of RWD, some limitations must be acknowledged. One major challenge is the variability in data quality and completeness. RWD are often collected from diverse sources, including electronic health records, insurance claims, and patient registries, which can lead to inconsistencies and gaps in the data. Additionally, the observational nature of RWD studies can introduce biases that are not present in randomized controlled trials. These biases can affect the validity of the findings and limit the generalizability of the results. Another limitation is the lack of standardization in data collection and reporting. Different healthcare systems and institutions may use varying methods to capture and record data, making it difficult to aggregate and compare data across different settings. Furthermore, the use of RWD requires sophisticated analytical techniques to account for confounding factors and ensure robust and reliable results. The consensus MDS and EDS proposed here will mitigate some of these limitations by helping participating clinics to collect standardized data. InRAD will provide an umbrella governance structure, including central statistical support to reduce bias by using advanced methods such as propensity score methods, instrumental variable analysis, and machine learning algorithms to control for confounding variables and improve the robustness of the results.

To ensure robust governance and ethical considerations for the InRAD registry, we draw inspiration from the MSBase registry, which provides a comprehensive framework for data ownership, participant privacy, and long-term sustainability. InRAD will be governed by a Global Board of Directors and a Scientific Leadership Group, ensuring strategic oversight and scientific integrity. Data ownership is clearly defined, with participating centers retaining ownership of their data while contributing to a centralized, de-identified dataset. Participant privacy is safeguarded through rigorous de-identification processes and adherence to ethical standards, including obtaining patient consent and securing ethics approval or exemption. Additionally, we emphasize long-term sustainability by operating as a not-for-profit organization, supported by a global network of specialist healthcare teams.

To ensure the global applicability of the InRAD registry, it is crucial to address regional disparities in access to biomarkers, imaging, and advanced therapies. These disparities can significantly impact the feasibility of data collection and the overall effectiveness of the registry. Therefore, we propose several strategies to mitigate these challenges. First, we recommend forming partnerships with local healthcare providers to facilitate access to necessary diagnostic and therapeutic resources. Second, we advocate for the implementation of standardized protocols that can be adapted to varying levels of available resources, ensuring consistency in data collection across diverse settings. Finally, we emphasize the importance of policy advocacy to improve access to advanced medical technologies in underserved regions. By proactively addressing these disparities, the InRAD registry aims to provide a comprehensive and equitable platform for real-world data collection in AD and other dementias, ultimately enhancing patient care and outcomes on a global scale.

## Conclusions

The primary aim was to define a MDS and EDS to collect RWD in InRAD and other AD registries. The MDS and EDS focus on informing routine clinical practice, covering relevant comorbidities and safety, and are designed to be easily integrated into existing data capture systems. The consensus MDS and EDS covers: patient profile and demographics; lifestyle and anthropometrics; co-morbidities and diagnostics; imaging; treatment; clinical characterization; safety; discontinuation; laboratory tests; patient and care partner outcomes; and interface functionality. The InRAD registry is being developed with the aim of advancing the field of AD care and research using RWD. By learning from successful examples in oncology, cardiology, diabetes, rheumatology and neuroinflammatory conditions like MS, addressing current limitations, and taking proactive steps to enhance data quality and analytical rigor, the registry will significantly contribute to improving patient care and outcomes in neurodegenerative diseases like AD.

The InRAD registry is designed to be a dynamic and evolving platform that will adapt to the rapidly advancing field of AD research. One of the key future directions for the registry is the integration of emerging biomarkers and therapies. As new biomarkers are identified, such as blood-based markers [73] and genetic markers [74], the registry will incorporate these into its data collection protocols. This will enhance the early detection and diagnosis of AD, allowing for more precise and personalized treatment approaches. In addition to biomarkers, the registry will also adapt to emerging therapies. Current research is exploring innovative treatments aimed at slowing disease progression and improving cognitive function. The InRAD registry will continuously update its data collection methods to capture information on these new treatments, ensuring that the registry remains at the forefront of AD research.

Furthermore, the registry will focus on expanding its global reach and inclusivity. This involves addressing regional disparities in access to diagnostic and therapeutic resources, as well as fostering collaborations with healthcare providers in underserved areas. By doing so, the InRAD registry aims to provide a comprehensive and equitable platform for real-world data collection, ultimately improving patient care and outcomes on a global scale.

## Data Availability

All data produced in the present study are available upon reasonable request to the authors

## Acknowledgements

Mark Greener, on behalf of TW1 Healthcare Consulting Limited, provided medical writing support. TW1 Healthcare Consulting Limited provided project management support. TW1 receives consulting fees from InRAD. RP is supported by the German Center for Neurodegenerative Diseases (Deutsches Zentrum für Neurodegenerative Erkrankungen, DZNE), the Hirnliga e.V. (Manfred-Strohscheer Stiftung) and the Deutsche Forschungsgemeinschaft (DFG, 1007 German Research Foundation) under Germany’s Excellence Strategy within the framework of 1008 the Munich Cluster for Systems Neurology (EXC 2145 SyNergy – ID 390857198), the Davos Alzheimer’s Collaborative, the VERUM Foundation, the Robert-Vogel-Foundation, the National Institute for Health and Care Research (NIHR) Sheffield Biomedical Research Centre (NIHR203321), the University of Cambridge – Ludwig-Maximilians-University Munich Strategic Partnership within the framework of the German Excellence Initiative and Excellence Strategy and the European Commission under the Innovative Health Initiative program (project 101132356).

The authors would like to thank the following for their contributions to the discussions regarding the MDS and EDS:

Carlos Acosta, Biogen

Jane Alty, University of Tasmania, Australia

Luisa Alves, Centro Clínico Académico de Lisboa-Nova Medical School, Portugal

Rhoda Au, Boston University, USA

Wing Chi Lisa Au, The Chinese University of Hong Kong, UK

Joanne Bell, Eisai

Başar Bilgiç, Istanbul University, Turkey

Vanessa Raymont, University of Oxford

Daniel J Blackburn, University of Sheffield, UK

Casper de Boer, Amsterdam UMC, the Netherlands

Riad Bournane, Eisai

Noa Bregman, Tel Aviv Medical Center, and Tel Aviv University, Israel

Amy Brodtmann, Monash University, Melbourne, Australia

Maja Katharina Grav Christensen, Eastern Health, Victoria, Australia

Sharon Cohen, Toronto Memory Program, Canada

Ana Sofia Costa, University Hospital RWTH, Aachen, Germany;

Elizabeth Coulthard, University of Bristol, UK

Virginie Dauphinot, Lyon University Hospital, France

Firuze Delen, Başakşehir Çam and Sakura City Hospital, Istanbul, Turkey

Sebastiaan Engelborghs, Vrije Universiteit Brussel and Universtair Ziekenhuis Brussels, Belgium

Nesrin Ergin, Pamukkale University, Denizli, Turkey

Maria Eriksdotter, Karolinska Institutet and Karolinska University Hospital, Stockholm, Sweden

Michael Ewers, University Hospital, LMU Munich, Germany

Ansgar Felbecker, Kantonsspital St Gallen, Switzerland

Tormod Fladby, Akershus University Hospital, University of Oslo, Norway

Kristian Steen Frederiksen, Rigshospitalet, Copenhagen, Denmark

Antoine Garnier-Crussard, Charpennes Hospital and Hospices Civils de Lyon, Villeurbanne, France

Hasmet Hanagasi, Istanbul Faculty Of Medicine, Turkey Masud Husain, University of Oxford, UK

Pervin Işeri, Yeni Yüzyıl University, Istanbul, Turkey

Jung Lung Hsu, New Taipei Municipal TuCheng Hospital, Taiwan

Ignacio Illán-Gala, Hospital de la Santa Creu i Sant Pau, Barcelona, Spain

Matthew Jones, Salford Royal Hospital, UK

Sean P Kennelly, Tallaght University Hospital, Dublin, Ireland

Chi-Hun Kim, Hallym University Sacred Heart Hospital, Pyeongchang, South Korea

Eun-Joo Kim, Pusan National University School of Medicine and Medical Research Institute, Busan, South Korea

Sean Knox, Biogen

Seong-Ho Koh, Hanyang University Guri Hospital, Guri-si, South Korea

Natasha Krishnadas, Florey Institute of Neurosciences & Mental Health, Victoria, Australia

Inês Laranjinha, Unidade Local de Saúde de Santo António, Porto, Portugal

Charlene Lee, Peninsula Health, Victoria, Australia

Teresa Leon, Novo Nordisk

Iracema Leroi, Trinity College and Global Brain Health Institute Dublin, Ireland

Johannes Levin, LMU Munich, Germany

Jae-Sung Lim, Asan Medical Center, Seoul, South Korea

Marco Lyons, Roche Products Ltd

Francesca Mangialasche, Karolinska Institutet, Stockholm, Sweden

Rafael Meyer, Psychiatrische Dienste Aargau AG, Windisch, Switzerland

Maas Christoph Mollenhauer, Wellington Hospital, New Zealand

Andreas U Monsch, University of Basel, Switzerland

So Young Moon, Ajou University School of Medicine, Suwon, South Korea

Diego Novick, Eli Lilly & Company

Sean O’Dowd, Tallaght University Hospital, Dublin, Ireland

Tiago Gil Oliveira, University of Minho, Portugal

Pierre Jean Ousset, Toulouse University Hospital, France

Alessandro Padovani, University of Brescia, Italy

Ming-Chyi Pai, Medical College and Hospital, National Cheng Kung University, Tainan, Taiwan

Richard J Perry, Imperial College and Imperial College Healthcare NHS Trust, London UK

Boris-Stephan Rauchmann, University Hospital LMU, Munich, Germany

Pascual Sánchez-Juan, Reina Sofia-CIEN Foundation-ISCIII, Madrid, Spain

Maria Isabel Jacinto Santana, Centro Hospitalar e Universit rio de Coimbra, Portugal

Nikolaos Scarmeas, National and Kapodistrian University of Athens, Greece and Columbia University, New York, USA

Jörg B. Schulz, RWTH Aachen University, Germany

Geir Selbaek, Vestfold Hositpal Trust, Tønsberg, Norway

Tamara Shiner, Tel Aviv University, Isreal

Cathy Short, Central Adelaide Local Health Network, Australia

Jón Snædal, Landspitali University Hospital, Reykjavík, Iceland

Eino Solje, University of Eastern Finland, Kuopio and Kuopio University Hospital Finland

Marc Sollberger, Die Universitäre Altersmedizin Felix Platter, Basel, Switzerland

Luiza Spiru, Ana Aslan International Foundation, Bucharest, Romania

Sofia Toniolo, University of Oxford, UK

Gorkem Tutal Gursoy, Ankara Bilkent City Hospital, Turkey

Sven J van der Lee, Amsterdam UMC, the Netherlands

Jo Vandercappellen, Eisai

Alberto Villarejo-Galende, Hospital Universitario 12 de Octubre and Universidad Complutense de Madrid, Spain

Huali Wang, Peking University Institute of Mental Health, China

Wendy Weidner, Alzheimer’s Disease International

Yuval Zabar, Biogen

