## Supplemental Table 1 for "Real-world datasets for the International Registry for Alzheimer’s Disease and Other Dementias (InRAD) and other registries: an international consensus"

**Supplemental table 1.** Domains identified by the International Steering Committee and items identified for both the minimum dataset and extended datasets

|  | **Minimum Data Set** | | **Extended Data Set** | |
| --- | --- | --- | --- | --- |
|  | **Domain** | **Frequency** | **Domain** | **Frequency** |
| **Patient profile and demographics** | Patient ID | Entry only | Other ID (for data linkage) | Entry only |
|  | Consent | Entry only | Care partner and patient consent for care partner to provide data | Entry and visits |
|  | Care partner and availability as informant | Entry and visits | Relationship to caregiver (spouse, partner, son or daughter, sibling, other relative, other [specify]) | Entry and visits |
|  | Sex assigned at birth | Entry only | Caregiver living with patient | Entry and visits |
|  | Birth year or date of birth | Entry only | Marital Status | Entry |
|  | Race or ethnicity | Entry only | Linguistics: Monolingual, bilingual or multilingual | Entry only |
|  | Country of birth and residence | Entry only | Dominant hand (left, right, ambidextrous) | Entry only |
|  | Education* | Entry only | Work status and current or last occupation | Entry and visits |
|  | Living status (alone, with family or partner, in care) | Entry and visits | Urbanicity of residence (city, town, rural) | Entry and visits |
| **Lifestyle and anthropometrics** | Height | Entry only | Alcohol consumption (units or standardized drinks per week) | Entry and visits |
|  | Weight | Entry and visits | Smoking (pack-years, current, ex-smoker, never smoker) | Entry and visits |
|  |  | | Cannabis and recreational drug use (drug[s], unknown, never, past, current) | Entry and visits |
|  |  |  | Physical activity (hours per week) | Entry and visits |
|  |  |  | Actively driving (date if stopped due to dementia) | Entry and visits |
|  |  |  | Sleep (hours per night; self and/or carer reports or using wearable sleep tracker) | Entry and visits |
| **Diagnostic work-up** | Family History of dementia first-degree relative/diagnosis | Entry only | Clinical specialty making the diagnosis | Entry only |
|  | Date of symptom onset | Entry only | Diagnosis Referral pathway (direct, primary care, other specialty)  Diagnosis specialty: (primary care, neurology, psychiatry, care of the elderly, other) | Entry only |
|  | Diagnosis (Alzheimer’s disease and/or co-morbidity or alternative diagnoses, including separate field for syndromic presentation without diagnosis) | Entry only | Diagnostic evidence of neurodegeneration Neurofilament light chain levels (yes, no, indeterminant, not performed) | Entry and visits |
|  | Predominant AD syndrome/variant in first 2 years (amnestic, posterior, logopenic or other) | Entry only | Previous or current dementia-related clinical trial (phase, dates, medication; entry only) | Entry and visits |
|  | Diagnostic Amyloid β status (yes, no, indeterminant, not performed) | Entry only |  | |
|  | Diagnostic Tau positivity status (yes, no, indeterminant, not performed) | Entry only |  |  |
|  | Diagnostic Imaging evidence of neurodegeneration (yes, no, indeterminant, not performed) | Entry only |  |  |
|  | Diagnostic imaging evidence of pathology secondary to AD (cerebral vascular lesions, cerebral amyloid angiopathy, other) | Entry only |  |  |
|  | Diagnostic biomarker evidence of neurodegeneration, such as neurofilament light and glial fibrillary acidic protein (yes, no, indeterminant, not performed) | Entry only |  |  |
|  | Relevant medical conditions** (history [entry only] and concomitant (entry and visits) | History: entry only  Concomitant: entry and visits |  |  |
| **Imaging** | Imaging (yes, no, date of scan) Type of scan (MRI/CT/PET/DAT) and reason (screening or diagnosis, monitoring progression and/or safety [eg ARIA]), other | Entry and visits | Scanner, magnet strength, Tracer used (eg fluorodeoxyglucose [FDG] Positron emission tomography) | When performed |
|  |  | | PET Quantification (for amyloid β and tau including post-treatment amyloid β quantification) (tracer and centiloid measure) | When performed |
|  |  |  | MRI/CT Atrophy (regional and global) and white matter lesion assessment and other findings (macro-hemorrhage; microhemorrhage; Superficial siderosis; Aria other) | When performed |
|  |  |  | Link to standard accepted minimum radiology report or quantification (yes, no) | When performed |
|  |  |  | Dopamine active transporter (DAT) scan | When performed |
|  |  |  | Laboratory amyloid β levels (CSF/Blood) | Entry and visits |
| **Laboratory tests (blood or cerebrospinal fluid)** | None | | Laboratory tau levels (CSF/Blood) | Entry and visits |
|  |  |  | Neurofilament light chain levels  GFAP | Entry and visits  Entry and visits |
|  |  |  | Biomaterial storage Y/N | Entry and visits |
|  |  |  | Genotype: apolipoprotein E status (note may be minimum for treatment screening) | Entry only |
|  |  |  | Genotype: other mutations and polymorphisms (*APP*, *PSY1*, *PSY2*) | Entry only |
|  |  |  | Complete blood count | When performed |
|  |  |  | Blood chemistry | When performed |
| **Treatment** | AD specific treatments (start/stop date, dose, unit, route and frequency) and reason for discontinuation or change ^¶^ | Entry and visits | Start/Change or stop date | Visits |
|  | Cognitive treatments of interest (entry and visits); including acetylcholinesterase inhibitors; partial antagonists of N-methyl-D-aspartate receptor (memantine); nutritional supplements; others | Entry and visits | Start/Change or stop date | Visits |
|  | Other treatments of interest^§^ | Entry and visits | Clinical Dementia Rating sum of boxes (CDR-SB) global score | Visits |
| **Clinical outcomes** | Global clinical staging (NIA-AA staging) | Entry and visits | Functional Activities Questionnaire (FAQ) score | When used |
|  | Cognitive screening test (yes, no; test MoCA or MMSE Score/version if applicable; | Entry and visits |  |  |
|  | Functional test (yes, no, name of test) | Entry and visits | Amsterdam Instrumental Activities of Daily Living Questionnaire (A-IADL-Q) score | When used |
|  | Milestone events other than Entry and visits  occupation and driving  (eg healthcare service use  and dependence); Has work,  driving and/or work status  changed since the last visit | | Neuropsychiatric Inventory Questionnaire (NPI*-*Q) | When used |
|  |  |  | Utilization in Dementia*-*Lite (RUD-Lite) score | When used |
|  |  |  | Medical events of interest other than those in the MDS (yes, no) | Visits only |
| **Safety** | Serious adverse event^†^ (Event since last visit yes, no and details including dates, MEDdra code, severity and outcome) | Visits only | Outcomes (eg hospitalization, out-patient, rehabilitation, treatment regimen modification) | Visits only |
|  | Amyloid-related imaging abnormalities (Event since last visit yes, no, not performed) | Visits only | Outcomes (eg hospitalization, out-patient, rehabilitation, treatment regimen modification) | When ARIA detected or DMT changed because of tolerability |
|  | Infusion/injection reactions (Event since last visit yes, no, details) | Visits only |  |  |
|  | Other medical events of interest^‡^ (Event since last visit yes, no, details) malignancy, non-ARIA related neurological conditions, serious infections | Visits only |  |  |
| **Patient and care partner outcomes** | None | | Quality of Life in Alzheimer's Disease scale (QoL*-*AD) score | When used |
|  |  |  | Euro-QoL 5 dimensions 5 levels (EQ-5D-5L) score | When used |
|  |  |  | Resource Utilization in Dementia*-*Lite (RUD-Lite) score | When used |
|  |  |  | Dependence Scale score | When used |
|  |  |  | Zarit Burden Interview score | When used |
|  |  |  | Neuropsychiatric Inventory Questionnaire (NPI*-*Q*)* | When used |
|  |  |  | Alzheimer's Disease Cooperative Study - Activities of Daily Living Scale for use in Mild Cognitive Impairment (ADSC_ADL-MCI) |  |
|  |  |  | After final visit | Clinical trial participation and/or eligibility |
| **Discontinuation** | Reason for discontinuation (including death) | After final visit |  |  |

* Using the International Standard Classification of Education [13].

** Cerebrovascular and other vascular or cardiac disorders, psychiatric conditions, other neurological/neurodegenerative conditions (including traumatic brain injury) as well as other significant other historic or on-going conditions including metabolic conditions (eg diabetes, obesity), malignancies, blood or lymphatic diseases, immune or autoimmune conditions, or history of severe or recurrent infections.

† Death, is life-threatening, permanent/serious disability or incapacity, requires/prolongs hospitalization, congenital anomaly/birth defect, other medical important condition [39].

‡ Including serious malignancy, serious infections, other non-ARIA related neurological conditions.

§ Antidepressants, anxiolytics, antipsychotics, antiseizure medications, sedatives, sleep aids (including continuous positive airway pressure), anti-diabetic and weight loss medication, cardiovascular medicines (eg antihypertensives, antilipidemic, antiplatelet, anticoagulants)

¶ Treatment-related adverse reaction or tolerability; lack of effectiveness or disease progression; patient or caregiver choice; scheduled stop or end of treatment; other (eg poor adherence) specified in a free text box.
